# Dysregulated anterior insula reactivity as robust functional biomarker for chronic pain – convergent evidence from neuroimaging meta-analysis

**DOI:** 10.1101/2021.03.24.21254023

**Authors:** Stefania Ferraro, Benjamin Klugah-Brown, Christopher R Tench, Shuxia Yao, Anna Nigri, Greta Demichelis, Maria Grazia Bruzzone, Benjamin Becker

## Abstract

Neurobiological pain models propose that the transition from acute to chronic pain is accompanied by neuropathological adaptations that mediate progressive pain processing dysfunctions. In contrast, meta-analytic studies on neurofunctional dysregulations in chronic pain have not revealed convergent evidence for robust alterations during experimental pain induction. Against this background, the present neuroimaging meta-analysis combined three different meta-analytic approaches with stringent study selection criteria for case-control functional magnetic resonance imaging experiments during acute pain processing with a focus on chronic pain disorders (i.e., fibromyalgia, irritable bowel syndrome, chronic low back pain, neuropathic pain; n = 295 patients, n = 211 controls; 86 foci). Across the meta-analytic approaches, convergent neurofunctional dysregulations in chronic pain patients were observed in the left anterior insula cortex, with study characteristics indicating generalized pain processing abnormalities. Seed-based resting-state functional connectivity based on a large publicly available dataset combined with a meta-analytic task-based approach identified the anterior insular region as a key node of an extended bilateral insula-fronto-cingular network, resembling the salience network. Moreover, the meta-analytic decoding showed that this region presents a high probability to be specifically activated during pain-related processes. Together, the present findings indicate that dysregulated left anterior insular activity represents a robust neurofunctional maladaptation and potential treatment target in chronic pain disorders.

## 1. Introduction

Chronic pain represents the major cause of disability and suffering across the world population (Goldberg and McGee, 2011; Rice et al., 2016). It has a tremendously detrimental impact on the physical and mental well-being of affected individuals, leading to considerable impairments in daily functioning and family lives (Dueñas et al., 2016). With prevalence rates as high as 10-30% in the adult population (Breivik et al., 2006; Reid et al., 2011), chronic pain leads to enormous social and economic burdens (Gaskin and Richard, 2012; Jackson et al., 2016).

According to the International Association for the Study of Pain (IASP), chronic pain refers to pain that lasts or recurs for at least 3 months (Treede et al., 2019). The IASP’s choice to focus on the duration of the symptoms was based on difficulties in operationalizing the previous definition, which emphasized functional differences between acute and chronic pain (”a pain persisting after the healing time and therefore that lacks its acute warning function”) (Treede et al., 2019). Unlike this recent pragmatic characterization of chronic pain for diagnostic purposes, neurobiological models emphasize that progressive (mal-)adaptive rearrangements of the brain at different scales (from molecules to networks) (Kuner and Flor, 2017) mediate the pathophysiological transition from acute to chronic pain.

Functional magnetic resonance imaging (fMRI) has dramatically advanced the comprehension of the neurobiological basis of acute and chronic pain in humans: there is now ample evidence that nociception engages the dynamic interplay among large-scale networks (Garcia-Larrea and Bastuji, 2018), with a recent neuroimaging meta-analysis suggesting that the experience of acute pain robustly engages a bilateral circuit encompassing somatosensory, insular, mid-cingulate, and thalamic regions (Xu et al., 2020a).

In contrast, neuroscientific research on the neural maladaptations that accompany the pathophysiological transition from acute into chronic pain remain to be clarified, although neurobiological mechanisms have been proposed (Vachon-Presseau et al., 2016; Denk et al., 2014; Tan et al., 2017) and a neural signature of chronic pain has been recently determined (Lee et al., 2021). Limitations inherent to the previous fMRI studies on chronic pain may have contributed to the lack of consistency, including a high proportion of false-positive and irreproducible results (Ioannidis, 2005) related to small sample sizes, variations in experimental designs and data processing pipelines, and insufficient control of the multiple comparisons problem (Poldrack et al., 2017; Eklund et al., 2016).

Quantitative meta-analyses, components of systematic reviews, represent the highest level of evidence and can overcome the limitations of the original studies by minimizing variability, systematic errors, and false positives. Several fMRI meta-analyses aimed at determining robust brain functional abnormalities in chronic pain conditions. Early investigations mainly employed the Activation Likelihood Estimation (ALE) algorithm (Turkeltaub et al., 2002; Eickhoff et al., 2009), however until 2016 (GingerALE version 2.3.5), this algorithm suffered from drawbacks in the code (Eickhoff et al., 2017) that, plausibly, invalidated several of these findings on chronic pain (Seifert and Maihöfner, 2011; Tillisch et al., 2011; Friebel et al., 2011; Simons et al., 2014; Jensen et al., 2016; Dehghan et al., 2016). Two more recent neuroimaging meta-analyses re-examined functional alterations in chronic pain patients during experimental pain processing utilizing either the updated version of the GingerALE algorithm (Xu et al., 2021) or a different algorithm (LocalALE in Tanasescu et al. (2016)). However, despite the great number of original studies (particularly in the work of Tanasescu et al. (2016)), both meta-analyses failed to determine convergent neurofunctional alterations in chronic pain patients during experimental pain induction. These negative findings stand in strong contrast with both neurobiological pain models (Tan et al., 2017; Vachon-Presseau et al., 2016) and clinical research demonstrating abnormal pain processing across chronic pain disorders (Staud and Domingo, 2001; Petersel et al., 2011; Chalaye et al., 2012; Goubert et al., 2017).

On this background, we performed a meta-analysis on fMRI studies to determine convergent neurofunctional alterations in chronic pain conditions during acute pain processing. In line with recent pathogenetic models suggesting that common neural maladaptations might mediate the transition into chronic pain (Vachon-Presseau et al., 2016), we expected to observe convergently dysregulated neural reactivity during acute pain infliction across chronic pain disorders. Considering that the negative findings in previous meta-analyses on chronic pain (Tanasescu et al., 2016; Xu et al., 2021) might be related to the high degree of heterogeneity of the selected studies, we adopted very stringent selection criteria to circumscribe this problem. First, in line with the recent meta-analysis from Xu et al. (2021), we selected only studies directly comparing chronic pain patients and healthy participants. This allowed us to avoid considerable heterogeneity introduced by different experimental settings, such as it occurs employing studies conducted separately in either pain patients or in healthy controls. Second, we focused on well-defined chronic pain conditions (e.g., fibromyalgia, irritable bowel syndrome, chronic low back pain, neuropathic pain) and excluded chronic pain conditions due to direct central diseases or insults such as brain tumors or brain trauma, which might be different from other chronic pain disorders in terms of the underlying neurobiological maladaptations.

To further promote a robust evaluation of convergent neurofunctional alterations, we employed two different coordinate-based meta-analytic algorithms (CBMA), namely GingerALE (Eickhoff et al., 2009, 2017) and the Coordinate Density Analysis (CDA) (Tench et al., 2020), and we functionally characterize the identified regions with a series of meta-analytic coactivation, decoding and network-level analyses.

## 2. Materials and Methods

### 2.1. Study selection

We followed the PRISMA reporting guidelines (Moher et al., 2009) for the present meta-analysis (see figure 1). Public databases of biomedical and life science literature reports, namely Pubmed, Web of Sciences, and Scopus, and the Neurosynth fMRI database (Yarkoni et al., 2011), were employed to search for fMRI or positron emission tomography (PET) studies investigating experimental cutaneous and visceral pain in four kinds of chronic pain conditions: neuropathic pain, fibromyalgia, irritable bowel syndrome, and low-back pain. The following keywords were used to search for papers present in the databases from inception to 26th July 2020: 1) irritable bowel syndrome: (“irritable bowel”OR “functional gastrointestinal”) AND (“functional magnetic resonance” OR “fMRI” OR “functional” OR “brain activation” OR “neural activity” OR “BOLD” OR “PET”); 2) fibromyalgia (“fibromyalgia” OR “fibrositis”) AND (“functional magnetic resonance” OR “fMRI” OR “functional” OR “brain activation” OR “neural activity” OR “BOLD” OR “PET”). 3. low-back pain (“low back pain” OR “chronic back pain” OR “chronic low back pain”) AND (“functional magnetic resonance” OR “fMRI” OR “functional” OR “brain activation” OR “neural activity” OR “BOLD” OR “PET”). 4. neuropathic pain (“neuropathic” OR “neuropathy”) AND (“functional magnetic resonance” OR “fMRI” OR “functional” OR “brain activation” OR “neural activity” OR “BOLD” OR “PET”). For each identified paper, we collected the title, author names, and date of publication. We merged the results from the different databases and identified and removed all duplicates. A unique identification number was assigned to the remaining papers. The abstracts were carefully read to exclude records investigating other disorders or not investigating brain activity. Then, reviewing the full text, we identified fMRI or PET studies investigating differences in brain activity between individuals suffering from chronic pain and control participants during experimental cutaneous or visceral noxious stimuli.

**Figure 1:**
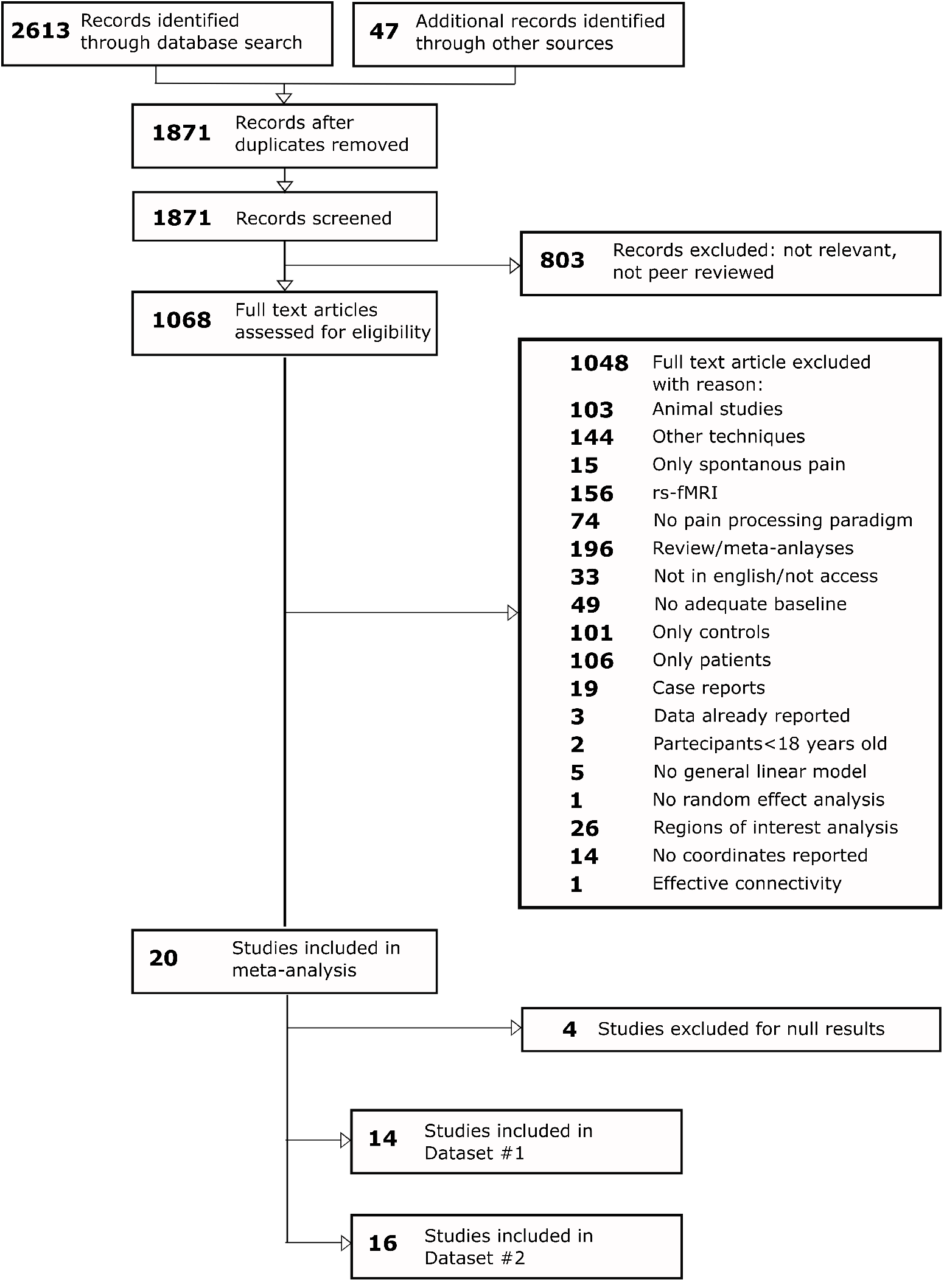
Flowchart of the screening process (PRISMA guideline)

We selected articles reporting between-group differences at a whole-brain level as coordinates in the standard Talairach Tournoux (TAL) (Talairach, 1988) or Montreal Neurological Institute (MNI) (Evans et al., 1993) stereotactic system. Moreover, only studies employing a general linear model approach and random effect analyses (Friston et al., 1999) were selected for this meta-analysis. Studies employing only regions of interest (ROIs) analyses were excluded, as well as studies investigating experimental pain processing only in patients or in control (CTRL) participants, or reporting only resting-state fMRI/PET results. Moreover, studies reporting brain activity during spontaneous pain in patients were not considered.

### 2.2 Data extraction

For each selected paper, we extracted the type of chronic pain condition investigated (neuropathic pain, fibromyalgia, irritable bowel syndrome, or low-back pain), the sample size, and the modality (i.e., visceral or cutaneous stimulation), the type (e.g., mechanical, thermal), and the body site of the administered nociceptive stimulation considering whether it was applied in a region where the chronic pain was experienced or not. Importantly, we also extracted whether opioid treatment was considered as an exclusion criterion for participation in the original study and whether patients discontinued or not their current pharmacological treatment before the MRI assessment. When not explicitly mentioned in the paper, we assumed that chronic pain individuals did not discontinue their medication for the scanning session. Since we were interested in the differences detected during the induction of experimental pain between chronic pain patients and control individuals, we also collected whether the two groups were matched for the same intensity of noxious stimulation or the same intensity of experienced pain. From the selected papers, one of the authors (S.F.) manually extracted the coordinates of the significant clusters resulting from the comparison between chronic pain patients and control individuals. Studies reporting no differences between groups were not considered for this meta-analysis. Standardized space (TAL or MNI) in which the peak coordinates were reported and the relative z-value or t-value were also extracted.

### 2.3. Quality control of the data

To evaluate the presence of possible coordinate extraction errors, three authors (G.D., A.N., MG.B.) independently checked the extracted coordinates. Then, an automatic diagnostic procedure implemented in NeuRoi and described in Tench et al. (2013) was employed. This tool computes a similarity score between the extracted coordinates of each selected study and a random selection of coordinates obtained from the same selected studies (Tench et al., 2013; Tanasescu et al., 2016). When potential errors were detected, coordinates were re-checked and corrected. All the analyses were conducted only after performing these steps.

### 2.4. Meta-analysis approaches: Ginger-ALE and Coordinate Density Analysis

There are multiple algorithms for performing CBMA, but each has different empirical parameters and assumptions, and each can produce different results conditional on the assumptions. Therefore, to obtain robust results, two different tools were employed: the GingerALE (Eickhoff et al., 2009) and the Coordinates Density Analysis (CDA) (Tench et al., 2020). With both methods, we detected common brain regions that showed abnormal brain activity in chronic pain patients in comparison to control individuals during experimental nociceptive stimulation. Importantly, GingerALE and CDA were employed to identify clusters of increased activity in chronic pain patients compared to control participants during experimental pain processing (Dataset #1: only coordinates of increased activity in chronic pain patients compared to control participants). Moreover, CDA was also employed to identify the clusters where chronic pain patients present increased and/or decreased activity in comparison to control participants (Dataset #2: coordinates of increased and/or decreased activity in chronic pain patients in comparison to control participants). We employed CDA to perform this analysis because it allows us to transparently interpret the results providing the data forest plot.

Due to the few investigations (n =8) showing decreased activity in chronic pain patients compared to control participants, we did not perform a separate analysis of these studies.

#### GingerALE

We employed GingerALE command-line version 3.0.2 (Eickhoff et al., 2009, 2012). It is well known that brains normalized to the MNI (Evans et al., 1993; Collins et al., 1994) and the Talairach (TAL) space (Talairach, 1988) present differences not only in size but also in origin, orientation (Lancaster et al., 2007), and shape (https://imaging.mrc-cbu.cam.ac.uk/imaging/MniTalairach). In particular, the brains in the MNI space are 5 mm taller and longer than the brains in the TAL space: therefore, they are larger and present lower temporal lobe and higher top than brains normalized to the TAL space (https://imaging.mrc-cbu.cam.ac.uk/imaging/MniTalairach).

This implies that the same coordinates in the two spaces do not refer to the same brain regions (Lancaster et al., 2007; Brett et al., 2002), posing issues in meta-analyses (Fox et al., 2005) in which studies conducted in different coordinate systems need to be analyzed together. Although the *icbm2tal* transformation matrix (Lancaster et al., 2007) was shown to be the most efficient algorithm in the corrections of the discrepancy between the two spaces (Laird et al., 2010), there is no definitive solution that completely solves this problem (https://imaging.mrccbu.cam.ac.uk/imaging/MniTalairach). To provide results not biased by the employed coordinates system (i.e., MNI or TAL), we conducted the coordinate-based analyses in both the MNI (Evans et al., 1993; Collins et al., 1994) and the TAL space (Talairach, 1988). As implemented in Ginger-ALE, *icbm2tal* transformation matrix was used to perform the coordinates conversions from one space to another. Therefore, for all the selected studies expressing the results in the MNI space, we implemented the conversion of the coordinates into the TAL space, while for all the selected studies expressing the results in the TAL space, we implemented the conversion in the MNI space. This produced two different foci formatting (i.e., one in the MNI space, the other in the TAL space).

For each selected study, the ALE algorithm, using a conservative mask and based on the number of the participants, computes the Full-Width Half-Maximum (FWHM) of the Gaussian kernel to smooth the identified foci and then creates the Modeled Activation Map (Eickhoff et al., 2009). These maps are then combined to obtain one single activation likelihood estimate (ALE) image, and normalized histograms are produced to create a table of p-values for the ALE scores. A permutation test (number of permutations = 1000), simulating random data, was used to define the statistical threshold to identify brain areas reported beyond the random chance (Eickhoff et al., 2009). In line with recent recommendations, we thresholded the ALE map at cluster-level inference threshold (cluster-level familywise error <0.05) and an uncorrected cluster forming threshold of p <0.001 (Eickhoff et al., 2012).

#### Coordinate density analysis (CDA)

CDA is a recently developed model-based method implemented in the NeuRoi image analysis software (https://www.nottingham.ac.uk/research/groups/clinicalneurology/neuroi.aspx), with very few apriori assumptions (Tench et al., 2020). This algorithm uses the density of coordinates from independent studies as its statistic and requires only one parameter: the human grey matter volume. Statistical thresholding to identify repeatable effects is performed by requiring a minimum proportion of the studies contributing to a cluster and is generally more conservative than false discovery rate (FDR at (q)=0.05); however, FDR is used as a limit on how liberal the threshold can get.

Importantly, this method, differently from Ginger-ALE, does not require the empirical choice of Gaussian smoothing kernel to extrapolate coordinates to voxel-wise activation maps or the randomization of the coordinates in the empirical space to define the statistical threshold (Tench et al., 2020). Employing CDA, the meta-analysis was conducted in the TAL space (Tench et al., 2020).

### 2.5. Functional characterization of the cluster of abnormalities

We conducted a functional characterization of any cluster emerging from the CDA performed on Dataset #2 (comprising the studies reporting increased and/or decreased activity in chronic pain patients compared to control participants). We took this decision because Dataset #2 presented the largest number of studies and participants and therefore an increased statistical power in comparison to Dataset #1.

To provide an unbiased functional characterization of clusters, we used Neurosynth, an open-source platform to synthesize large-scale fMRI data (http://neurosynth.org) (Yarkoni et al., 2011), which presents, to date, 14371 fMRI studies for a total of 507891 activations. Each article in the Neurosynth database is automatically text-mined to identify specific terms occurring with high-frequency, and then it is “tagged” with these terms. The activation coordinates are automatically extracted, and meta-analyses can be performed (Yarkoni et al., 2011). Before any analyses, the TAL coordinates of the observed cluster were transferred in the MNI space employing the GingerALE algorithm since Neurosynth works in this space.

#### Seed-based functional connectivity

In an attempt to identify the neural circuit that might be most likely abnormal in patients with chronic pain, we produced a conjunction map of the resting-state functional connectivity (rs-FC) map and of the meta-analytic coactivation map, both seeded in the peak coordinates of clusters of interest and both generated with the Neurosynth function *locations*. This allowed us to obtain a robust condition-independent (i.e., independent from “task free” and “task-based” states) identification of the functional network connected to the cluster of interest (Schnellbächer et al., 2020; Rottschy et al., 2013).

The seed-based rs-FC map was obtained from a large rs-fMRI dataset (1000 human subjects), which results are provided in Neurosynth as a courtesy of T. Yeo, R. Buckner, and the Brain Genomics Superstructure Project (Buckner et al., 2011; Yeo et al., 2011). The corresponding Pearson’s correlation coefficients (r) maps express the strength of the correlation between the time-series rs-fMRI activity in the seed (∼4×4 mm vertex, but to be considered more extended, see Yeo et al. (2011) for details) and the activity observed in each brain vertex during “task-free” condition. This uncorrected map was thresholded at r>0.2 (Yeo et al., 2011).

The seed-based meta-analytic coactivation map was obtained from an automatic meta-analysis performed on the Neurosynth platform across all the fMRI studies present in the database. This z-score map (FDR-corrected, (q)=0.01) shows the degree of the association between the presence/absence of the activity in the 6mm spherical seed and the presence/absence of the activity observed in each voxel during “task-based” conditions (https://www.neurosynth.org/locations/).

Then, we produced a conjunction map of the seed-based rs-FC map and of the seed-based meta-analytic coactivation map (Schnellbächer et al., 2020; Rottschy et al., 2013). To this aim, we first converted the rs-FC r-values map in z-scores map employing the inverse Fisher’s r-to-z transformation, and then, to compute the conjunction map, we employed the minimum statistics approach (Jakobs et al., 2012; Nichols et al., 2005). The probabilistic cytoarchitectonic atlas implemented in SPM Anatomy Toolbox (Eickhoff et al., 2005) was used to label the regions larger than 50 voxels belonging to this network.

#### Decoding

Using NiMARE python packages (https://nimare.readthedocs.io/en/latest/), we decoded the 6mm spherical region of interest (ROI) centered in the coordinate of the cluster that emerged from the meta-analysis. The decoding quantifies the probability that a specific brain region is activated given the presence of a specific term ([*P*(*activation* | *term*)], or forward inference) and the probability that a specific term is present given the activity in a specific brain area ([*P*(*term* | *activation*)], or ‘reverse inference’). Therefore, based on the assumption that terms present in a work are as proxies for cognitive processes, the forward inference allows to probabilistically identify the regions that are consistently reported with a certain cognitive process, while and more importantly, the reverse inference allows to probabilistically identify the cognitive process that might be selectively associated with the activity of a particular region (Poldrack, 2011; Yarkoni et al., 2011).

To this aim, we produced a 6mm spherical region of interest (ROI) centered in the coordinate of the cluster of interest, and then we applied the function *nimare.decoder.discrete* on the whole Neurosynth database. From the decoder’s output, we arbitrarily decided to report the first 15 terms ordered in descending order according to the conditional probability of the reverse inference [*P*(*term* | *activation*)]. From this list, we excluded terms reporting anatomical regions and generic terms (such as “networks involved”, “nervous”, “spatial-temporal”). As a post-hoc analysis, based on the hypothesis that the cluster identified with the meta-analysis might also be involved in interoceptive functions, we decoded the terms “autonomic” and “interoceptive” employing the Neurosynth *term-based meta-analytic* function.

## 3. Results

### 3.1. Study sample characteristic

Based on the selection criteria 1, we identified 14 studies (Dataset #1) reporting increased activity in chronic pain patients in comparison to control participants (for details, see table 1 and 2). These 14 studies investigated a total of 263 chronic pain patients (chronic pain patients in each study, mean: 19, median:16, range: 8-48) and 179 control subjects (mean:13, median:11, range:8-21). These studies investigated fibromyalgia (5, 36% of studies), irritable bowel syndrome (4, 29% of studies), neuropathic pain (2, 14% of studies), low-back pain (2, 14% of studies) and neuropathic pain and fibromyalgia (1, 7%). In this dataset, 64 total foci were identified: fibromyalgia and irritable bowel syndrome studies contributed respectively for 52% and 30% of the total foci, while neuropathic pain and low back pain contributed respectively for 14% and 3%, the study conjointly investigating neuropathic pain and fibromyalgia contributed for 2%.

**Table 1:** Studies characteristics.* Studies contributing to the cluster determined in the analysis of Dataset #2 employing the coordinate density analysis (CDA) (Tench et al., 2020). Abbreviations: CP = chronic pain patients, CTRL = control individuals, F = female, M = male.

| Studies | Participants(#) |  | Duration<br>chronic pain<br>(months) | Between-group differences direc-<br>tion for scores of: |  | Medications wash-out<br>(yes/no); time before<br>scanning session |
| --- | --- | --- | --- | --- | --- | --- |
|  | CP(F/M) | CTRL(F/M) |  | Depression | Anxiety |  |
| Fibromyalgia |  |  |  |  |  |  |
| Gracely et al. (2002) | 16(15/1) | 16(15/1) | Not declared | Not declared | Not declared | Yes; 12 hours |
| Pujol et al. (2009)* | 9(9/0) | 9(9/0) | 104 (M) | Not declared | Not declared | Yes; 72 hours |
| Jensen et al. (2009) | 16(16/0) | 16(16/0) | Not declared | Not declared | Not declared | Yes; flexible |
| Burgmer et al. (2012) | 17(17/0) | 17(17/0) | >24 | CP>CTRL | CP>CTRL | Yes; 48 hours |
| Kim et al. (2013) | 21(21/0) | 11(11/0) | Not declared | CP>CTRL | CP=CTRL | Not declared |
| Schreiber et al. (2017) | 38(33/5) | 15(10/5) | Not declared | CP >CTRL | CP>CTRL | No wash-out |
| Low-back pain |  |  |  |  |  |  |
| Derbyshire et al. (2002)* | 16(12/4) | 16(11/5) | Not declared | trend CP>CTRL | CP>CTRL | Yes; 2 weeks |
| Baliki et al. (2010) | 16(8/8) | 16(8/8) | Not declared | Not declared. | Not declared | Not declared |
| Rodriguez-Raecke et al. (2014) | 19(12/7) | 21(15/6) | >2 | CP>CTRL | CP=CTRL | No wash-out |
| Irritable bowel syndrome |  |  |  |  |  |  |
| Verne et al. (2003)* | 9(6/3) | 9(6/3) | >60 | Not declared | Not declared | Not declared |
| Elsenbruch et al. (2010)* | 15(15/0) | 12(12/0) | >12 | CP=CTRL | CP>CTRL | Not declared |
| Bouhassira et al. (2013)* | 19(19/0) | 11(11/0) | 216 | CP=CTRL | CP=CTRL | Yes; 7 days |
| Guleria et al. (2017)* | 20(0/20) | 10(0/10) | 28 | Not declared | Not declared | Not declared |
| Neuropathic pain |  |  |  |  |  |  |
| Albuquerque et al. (2006) | 8(8/0) | 8(8/0) | 48 | CP = CTRL | CP>CTRL | Yes; >4 half-lives |
| Vartiainen et al. (2009)* | 8(7/1) | 11(5/6) | 36–240 | Not declared | Not declared | Not declared |
| Neuropathic& fibromyalgia |  |  |  |  |  |  |
| Hampson et al. (2013)* | 48(48/0) | 13(13/0) | Not declared | Not declared | Not declared | Not declared |
| Total participants | 295(246/49) | 211(167/44) |  |  |  |  |

**Table 2:** Studies characteristics.* Studies contributing to the cluster determined in the analysis of Dataset#2 employing the coordinate density analysis (CDA). *Foci(#): CPvs. CTRL*: number of foci of abnormal activity (↑ = increased activity, ↓= decreased activity) emerged in the comparison between chronic pain patients and control participants. *Stimulus modality* = type of painful stimulation applied for inducing pain; *Stimulation site (In/Out chronic pain body regions)*: part of the body subjected to painful stimulation and whether this body part was located in or out the regions where chronic pain is normally experienced; *Groups matching*: the group of chronic pain patients and of control participants were matched for the experienced pain (*Pain*) or for the intensity of painful stimulus (*Stimulus*); *Pain scores or Stimulus intensity*: scores on VAS or Likert scale for the perceived pain (when groups were matched for perceived pain) or intensity of the administered stimulation (when groups were matched for the intensity of painful stimulation); *Direction of the between-group difference for pain(P) or stimulation (S)*: direction of the behavioural differences in the perceived pain (when groups were matched for the intensity of painful stimulation) or in the intensity of the applied painful stimulation (when groups were matched for perceived pain). Abbreviations: CP = chronic pain patients, CTRL = control individuals, L = left, R = right, press = pressure; dist = distension; mod-sev = from moderate to severe.

| Studies | Foci(#): CP vs. CTRL |  | Stimulation modality | Stimulation site (in/out chronic pain body regions) | Groups matching (for perceived pain or stimulus intensity) | Pain scores or stimulation intensity | Direction of the between-group difference for pain (P) or stimulation (S) | Validated scale for diagnosis |
| --- | --- | --- | --- | --- | --- | --- | --- | --- |
|  | ↑ | ↓ |  |  |  |  |  |  |
| Fibromyalgia |  |  |  |  |  |  |  |  |
| Gracely et al. (2002) | 13 | 1 | Mechanical (press) | L thumb (Out) | Stimulus | 0.45-4.5 kg/mm<sup>2</sup> | CP>CTRL(P) | American College Rheumatology |
| Pujol et al. (2009)* | 5 | 0 | Mechanical (press) | R thumb (Out) | Pain | 70/100 | CP<CTRL(S) | American College Rheumatology |
| Jensen et al. (2009) | 0 | 2 | Mechanical (press) | Thumb (Out) | Pain | 50/100 | CP<CTRL(S) | Not declared |
| Burgmer et al. (2012) | 1 | 2 | Mechanical (press) | R forearm (Out) | Stimulus | Incision | CP>CTRL(P) | American College Rheumatology |
| Kim et al. (2013) | 8 | 0 | Mechanical (press) | L thumb (Out) | Pain | 14/21 | CP<CTRL(S) | Not declared |
| Schreiber et al. (2017) | 6 | 0 | Mechanical (press) | L calf (Out) | Pain | 40/100 | CP<CTRL(S) | American College Rheumatology |
| Low-back pain |  |  |  |  |  |  |  |  |
| Derbyshire et al. (2002)* | 1 | 3 | Thermal | R hand (Out) | Pain | 40-60/100 | CP= CTRL(S) | Not declared |
| Baliki et al. (2010) | 0 | 2 | Thermal | Back (In) | Stimulus | 47-51° | CP= CTRL(P) | Not declared |
| Rodriguez-Raecke et al. (2014) | 1 | 0 | Thermal | L forearm (Out) | Stimulus | 48°C | CP= CTRL(P) | Quebec Task Force |
| Irritable bowel syndrome |  |  |  |  |  |  |  |  |
| Verne et al. (2003)* | 8 | 2 | Mechanical (dist) | Rectum (In) | Stimulus | 55mmHg | CP>CTRL(P) | Rome III |
| Elsenbruch et al. (2010)* | 2 | 0 | Mechanical (dist) | Rectum (In) | Pain | 5/6 | CP=CTRL(S) | Rome III |
| Bouhassira et al. (2013) * | 5 | 0 | Mechanical (dist) | Rectum (In) | Pain | 5/6 | CP <CTRL(S) | Rome III |
| Guleria et al. (2017)* | 4 | 2 | Mechanical (dist) | Rectum (In) | Pain | mod-sev | CP <CTRL(S) | Rome III |
| Neuropathic pain |  |  |  |  |  |  |  |  |
| Albuquerque et al. (2006) | 6 | 3 | Thermal | R trigeminal (In) | Stimulus | 47-49°C | CP=CTRL(P) | Not declared |
| Vartiainen et al. (2009)* | 3 | 7 | Thermal | Bilat. hand (Out) | Pain | 6-7/10 | CP<CTRL(S) | Not declared |
| Neuropathic& fibromyalgia |  |  |  |  |  |  |  |  |
| Hampson et al. (2013)* | 1 | 0 | Mechanical | L thumb (Out) | Pain | 13.5/21 | CP<CTRL(S) | Not declared |
| Total foci | 64 | 22 |  |  |  |  |  |  |

Moreover, we identified a second dataset (Dataset #2) (for details, see table 1 and 2), which included coordinates of increased as well as decreased activity in chronic pain patients relative to controls. This dataset comprised all studies from Dataset #1, as well as two additional investigations reporting only decreased activity (investigating fibromyalgia and low-back pain). These 16 studies investigated a total of 295 chronic pain patients (mean: 18; median: 16; range:8-48) and 211 control subjects (mean: 13; median:12; range:8-21). From this dataset, we identified 86 total foci. Studies investigating fibromyalgia and irritable bowel syndrome contributed respectively for 44% and 24% of the total foci, while studies investigating neuropathic pain and low back pain contributed respectively for 22% and 8% of the total foci, the study conjointly investigating neuropathic pain and fibromyalgia contributed for 1%.

Across Dataset #1 experimental nociception was induced by mechanical cutaneous stimulation (5 studies on fibromyalgia and 1 study on neuropathic pain fibromyalgia), by mechanical visceral stimulation (4 studies on irritable bowel syndrome), and by thermal cutaneous stimulation (2 studies on neuropathic pain, 2 studies on low-back pain). Dataset #1 comprised 5 studies employing the same intensity of painful stimulation and 9 studies employing the same level of perceived pain to test for differences between the group of chronic pain patients and control participants.

Similarly, in Dataset #2, experimental nociception was induced by mechanical cutaneous stimulation (6 studies on fibromyalgia and 1 study on neuropathic pain fibromyalgia), by mechanical visceral stimulation (4 studies on irritable bowel syndrome), and by thermal cutaneous stimulation (2 studies on neuropathic pain, 3 studies on low-back pain). Moreover, 6 studies employed the same intensity of painful stimulation while 10 studies employed the same level of perceived pain to match the chronic pain patients group and the control group. Importantly, 6 studies (all the 4 studies investigating irritable bowel syndrome, 1 study investigating neuropathic pain, and 1 investigating low-back pain) administered the experimental painful stimulation in the region where chronic pain was experienced. The medication wash-out, with variable temporal windows, was employed in 7 studies, while 2 declared that patients were not asked to discontinue the assumption of their medications. The remaining studies did not explicitly report medication status, therefore, we assumed that the patients did not discontinue their usual medications. Notably, in both datasets, there was a higher proportion of women (Dataset #1: total chronic pain patients = 263; 222 women, 84% of the sample; Dataset #2: total chronic pain patients: 295; 246 women: 83% of the sample).

### 3.2. Ginger-ALE results

The ginger-ALE analyses employing the two different coordinate systems (TAL and MNI space) on Dataset #1 provided convergent results revealing the same 2 clusters (formed by 6 studies in total) of higher fMRI activation in chronic pain patients in comparison to control participants. The first cluster was detected in the left anterior insula cortex, and it was centered at [−36.8,15.2,3.2] in TAL space and at [−38.7,17.9,−2.1] in MNI space. When the coordinates obtained in the TAL space [−36.8,15.2,3.2] were converted to MNI space using the algorithm implemented in GingerALE, results resembled the results as by MNI space analysis (i.e. [−38.7, 17.9, −2.1]). In both analyses, 5 studies contributed to this cluster: all 4 studies investigating irritable bowel syndrome, and 1 investigating fibromyalgia. The second cluster of increased activity emerged in both analyses in the left claustrum/anterior insula cortex, and it was centered at [−31.1,0.8,9.6] in TAL space and at [−32.4,3.3,6.4] in MNI space. Three studies (2 of them also contributing to the first cluster) contributed to this second cluster (1 study on neuropathic pain, 1 on irritable bowel syndrome, and 1 on neuropathic pain and fibromyalgia) (see table 3).

**Table 3:** Studies and the relative foci expressed as coordinates in the Talairaich space contributing to the identified clusters employing the Coordinate Density Analysis (CDA) (Tench et al., 2020) and GingerALE (Eickhoff et al., 2009, 2017) on Dataset #1 and Dataset #2.

| **CDA** |  |  |  |  |  | **GingerALE** |  |  |  |  |
| --- | --- | --- | --- | --- | --- | --- | --- | --- | --- | --- |
| **Dataset #1** | Contributors to cluster #1 [-36 12 4] |  |  |  |  | Contributors to cluster #1 [-36.8, 15.2, 3.2] |  |  |  |  |
|  | **Studies** | x | y | z | z-score | **Studies** | x | y | z | z-score |
|  | Bouhassira et al. (2013) | -32.8 | 15.3 | -1.7 | 5.8 | Bouhassira et al. (2013) | -32.8 | 15.3 | -1.7 | 5.8 |
|  | Elsenbruch et al. (2010) | -36.0 | 18.0 | 1.0 | 3.6 | Elsenbruch et al. (2010) | -36.0 | 18.0 | 1.0 | 3.6 |
|  | Guleira et al. (2017) | -40.3 | 11.7 | 7.9 | 4.0 | Guleira et al. (2017) | -40.3 | 11.7 | 7.9 | 4.0 |
|  | Verne et al. (2013) | -35.0 | 18.0 | 4.0 | 2.7 | Verne et al. (2013) | -35.0 | 18.0 | 4.0 | 2.7 |
|  | Pujol et al. (2009) | -42.0 | 12.0 | 5.0 | 3.6 | Pujol et al. (2009) | -42.0 | 12.0 | 5.0 | 3.6 |
|  | Hampson et al. (2013) | -34.8 | -1.7 | 10.5 | 4.0 |  |  |  |  |  |
|  |  |  |  |  |  | Contributors to cluster #2 [-31.1, 0.8, 9.6] |  |  |  |  |
|  |  |  |  |  |  | Hampson et al. (2013) | -34.8 | -1.7 | 10.5 | 4.0 |
|  |  |  |  |  |  | Verne et al. (2013) | -31.0 | 2.0 | 10.0 | 2.8 |
|  |  |  |  |  |  | Pujol et al. (2009) | -27.0 | 3.0 | 8.0 | 2.7 |
| **Dataset #2** | Contributors to cluster #1 [-34 14 6] |  |  |  |  |  |  |  |  |  |
|  | **Studies** | x | y | z | z-score |  |  |  |  |  |
|  | Bouhassira et al. (2013) | -32.8 | 15.3 | -1.7 | 5.8 |  |  |  |  |  |
|  | Elsenbruch et al. (2010) | -36.0 | 18.0 | 1.0 | 3.6 |  |  |  |  |  |
|  | Guleira et al. (2017) | -40.3 | 11.7 | 7.9 | 4.0 |  |  |  |  |  |
|  | Verne et al. (2013) | -35.0 | 18.0 | 4.0 | 2.7 |  |  |  |  |  |
|  | Pujol et al. (2009) | -42.0 | 12.0 | 5.0 | 3.6 |  |  |  |  |  |
|  | Hampson et al. (2013) | -34.8 | -1.7 | 10.5 | 4.0 |  |  |  |  |  |
|  | Derbyshire et al. (2002) | -31.0 | 15.4 | 8.3 | -2.8 |  |  |  |  |  |
|  | Vartiainen et al. (2009) | -31.0 | 14.0 | 13.0 | -3.8 |  |  |  |  |  |

Importantly, among the studies contributing to the observed clusters, 5 matched the patient and control groups for the intensity of perceived pain (3 studies on irritable bowel syndrome, 1 on fibromyalgia, and 1 on neuropathic pain and fibromyalgia), while 1 for the intensity of stimulation (1 on irritable bowel syndrome). These investigations employed visceral mechanical simulation (4 studies on irritable bowel syndrome) and cutaneous mechanical stimulation (1 study on fibromyalgia and 1 on neuropathic pain and fibromyalgia). All the irritable bowel syndrome studies employed the nociceptive stimulation in the domain where chronic pain was experienced (rectum), while the remaining 2 studies employed a mechanical stimulation outside the domain where chronic pain was experienced.

### 3.3. Coordinate density analysis results

CDA results of Dataset #1 revealed a single significant cluster (minimum number of studies = 5) of increased activity in the left anterior insula cortex centered on [−36, 12, 4] TAL space. Notably, the coordinates of this cluster were very similar to the coordinates of the first cluster emerged in the ALE approach [−36.8,15.2,3.2]. Six investigations contributed to this cluster: 5 of these also contributed to the respective cluster obtained with GingerALE.

The CDA results of Dataset #2 (see fig. 2) again revealed a cluster of significant functional abnormalities in the left anterior insula (centered in [−34, 14, 6] TAL space) (minimum number of studies = 5) (figure 2).

**Figure 2:**
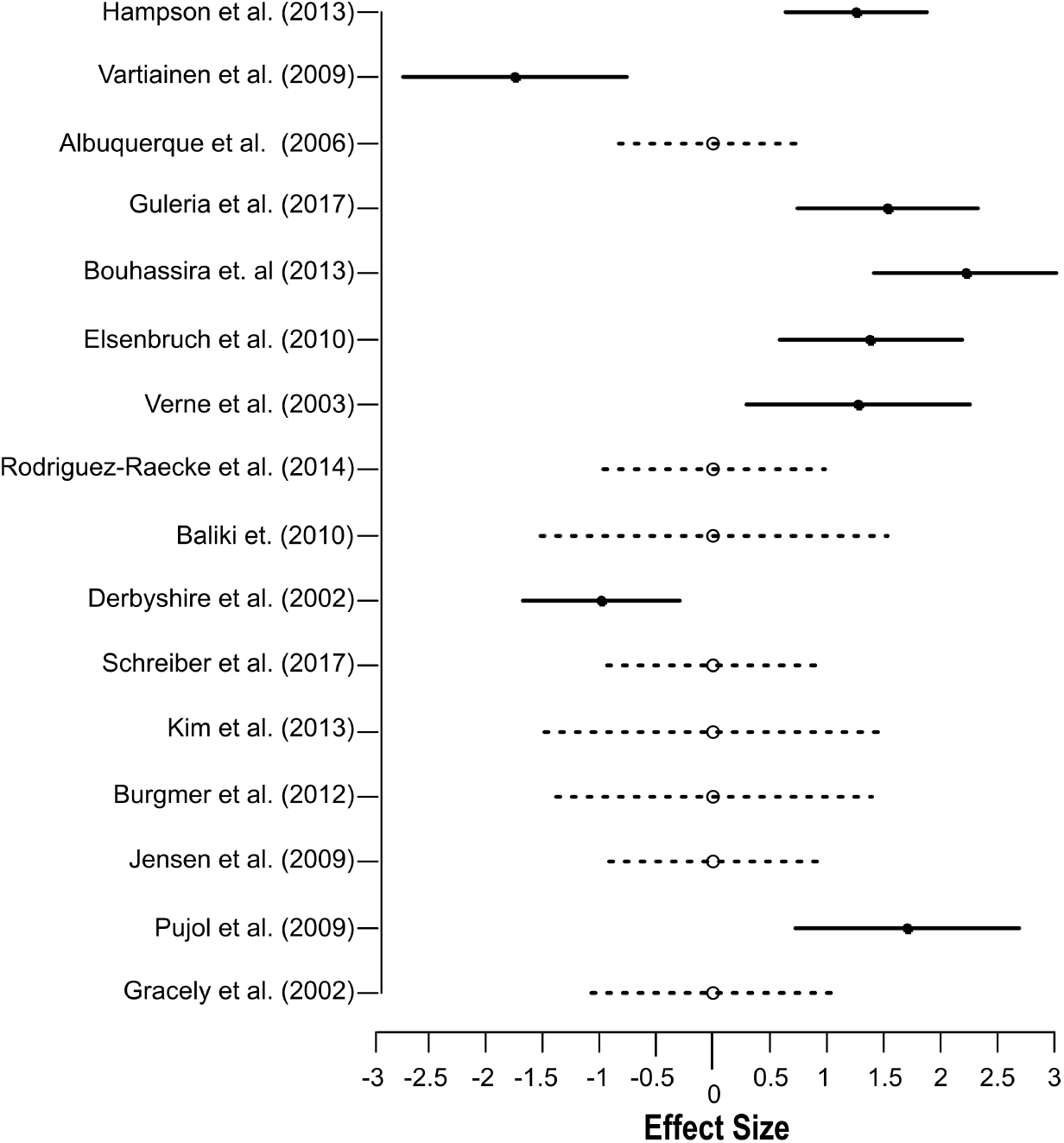
Effect size of the studies contributing to the cluster of abnormality emerged from the coordinate density analysis (CDA) (Tench et al., 2020) of the Dataset2 (comprising studies showing increased and/or decreased activity in chronic pain patients in comparison to control participants.)

**Figure 3:**
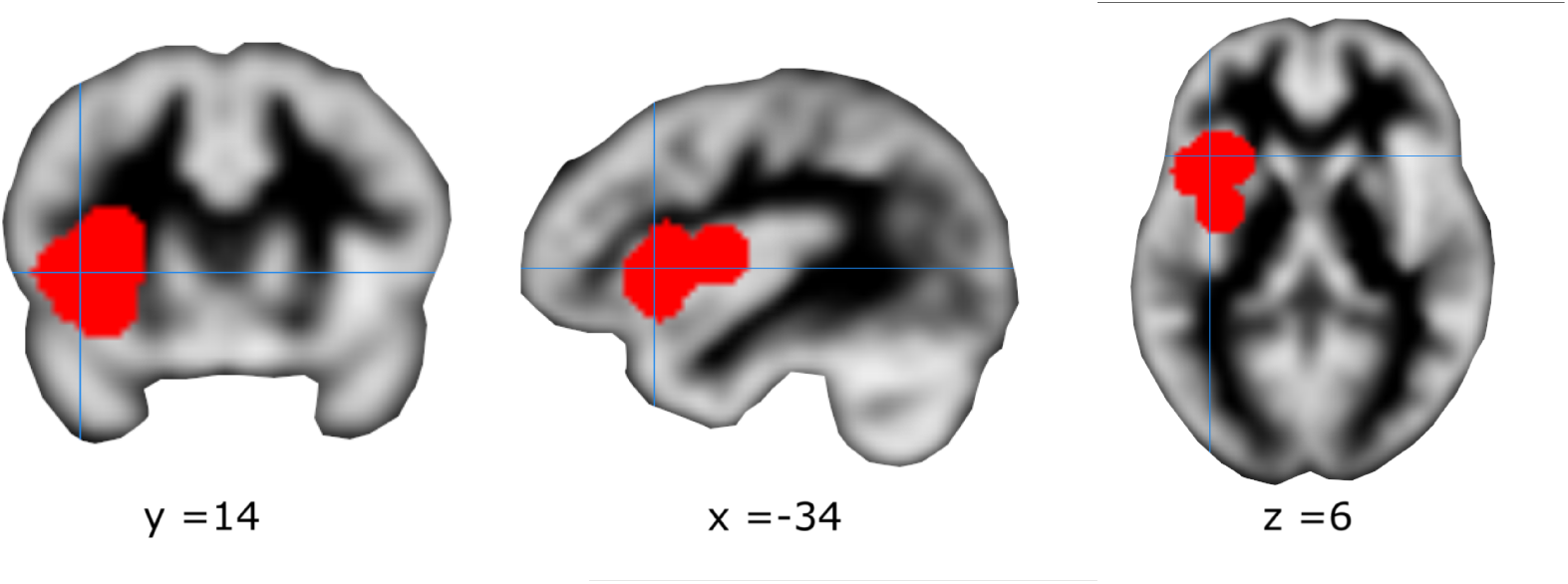
Cluster of convergent activity emerged from the coordinate density analysis (CDA) on Dataset #2. Mean of the coordinates of the cluster in the Talairach space [−34, 14, 6].

Notably, in this analysis, 8 studies contributed to this cluster: 4 investigating irritable bowel syndrome, 1 fibromyalgia, 1 low-back pain, 1 neuropathic pain, and 1 fibromyalgia and neuropathic pain. As expected, this cluster was formed by all the studies contributing to the cluster obtained with Dataset #1 (see table 3) and two more additional studies (1 on neuropathic pain and 1 on chronic low-back pain). Importantly, 7 studies contributing to the observed cluster matched the patient and control groups for the intensity of perceived pain and only 1 study for the intensity of stimulation (1 study on irritable bowel syndrome).

### 3.4. Functional characterization

#### Seed-based functional connectivity

The conjunction map obtained from the rs-FC map and the meta-analytic co-activation map both seeded in the region of altered neural activity in chronic pain patients (CDA on Dataset #2; MNI coordinate: [−36, 17, 1]), revealed a widespread bilateral network (table 4 and figure 4). Specifically, three large connected clusters were identified, the first encompassing the right insula lobe and the right inferior and middle frontal gyri, the second encompassing the left insula lobe, the left inferior frontal regions (inferior frontal gyrus, the Rolandic operculum), and the left precentral gyrus, while the third encompassed the right midcingulate cortex, the left anterior cingulate cortex, and the left posterior medial frontal gyrus. Additional clusters of this functional network were also found in the left inferior and middle frontal gyrus, bilateral parietal regions (supramarginal gyrus and inferior parietal lobule), and bilateral globus pallidum.

**Table 4:** Anatomical regions and MNI coordinates of the peaks of the conjunction analysis performed with the minimum statistic approach (Jakobs et al., 2012; Nichols et al., 2005) between the resting-state functional connectivity (rs-FC) map and the meta-analytic coactivation map both seeded in the MNI coordinates[−36 17 1] of the significant cluster emerged from the coordinate density analysis (CDA) on Dataset #2. Regions larger than 50 voxels were automatically labeled with Anatomy toolbox (Eickhoff et al., 2005). Abbreviations: Lat. = hemisphere, L= left, R = right, K_*E*_ = number of voxels.

| | $K_E$ | MNI coord. | | | Lat. | Anatomical region |
| --- | --- | --- | --- | --- | --- | --- |
|  |  | x | y | z |  |  |
| Cluster #1 | 3691 | 42 | 24 | 8 | R | Insula lobe |
|  |  | 36 | 50 | 20 | R | Middle frontal gyrus |
|  |  | 52 | 12 | 24 | R | Inferior frontal gyrus |
| Cluster #2 | 2563 | -36 | 16 | 0 | L | Insula lobe |
|  |  | -50 | 10 | 8 | L | Inferior frontal gyrus |
|  |  | -52 | 6 | 14 | L | Precentral gyrus |
|  |  | -40 | -4 | 12 | L | Rolandic operculum |
| Cluster #3 | 2439 | 2 | 18 | 44 | R | Posterior-medial frontal gyrus |
|  |  | 2 | 20 | 40 | R | Midcingulate cortex |
|  |  | 2 | 34 | 24 | L | Anterior cingulate cortex |
|  |  | 4 | 2 | 62 | R | Posterior-medial frontal gyrus |
| Cluster #4 | 1399 | -38 | 46 | 18 | L | Middle frontal gyrus |
|  |  | -42 | 32 | 20 | L | Inferior frontal gyrus |
| Cluster #5 | 852 | -40 | -46 | 42 | L | Inferior parietal Lobule |
|  |  | -60 | -34 | 36 | L | Supramarginal gyrus |
| Cluster #6 | 709 | 38 | -44 | 44 | R | Inferior parietal lobule |
|  |  | 52 | -38 | 44 | R | Supramarginal gyrus |
| Cluster #7 | 94 | 14 | 10 | 4 | R | Pallidum |
| Cluster #8 | 86 | -12 | 6 | 4 | L | Pallidum |

**Figure 4:**
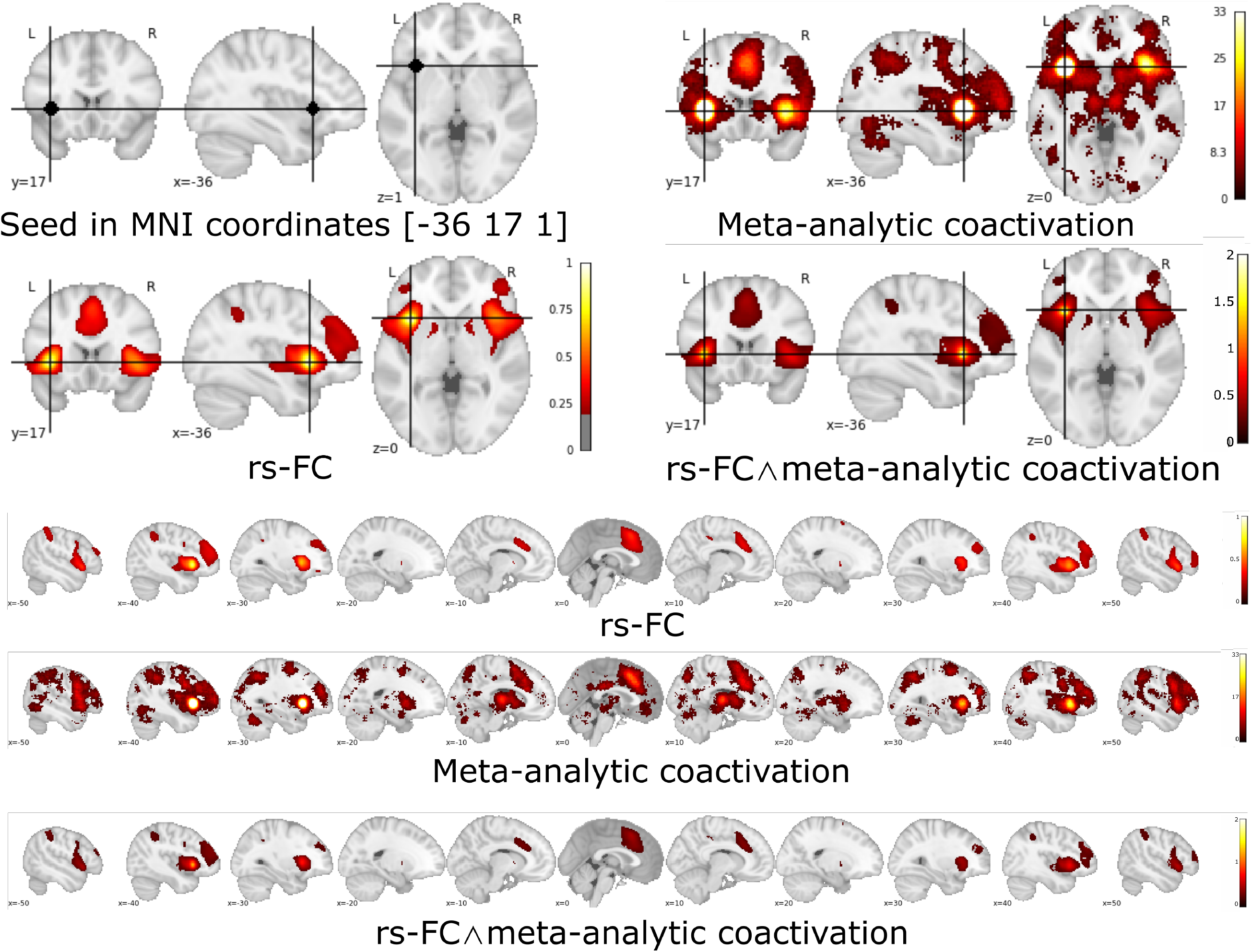
Seed-based functional connectivity analyses (seed in the mean of the coordinates of the cluster obtained from the coordinate density analysis (CDA) on Dataset #2): resting-state functional connectivity (rs-FC) map, meta-analytic coactivation map, and the relative conjunction map [rs-FC map Coactivation map]. Projection on coronal, sagittal and transversal plane and multislice on the sagittal plane.

#### Decoding

Decoding the functional properties of the identified left anterior insula region showed that among the first 15 terms ordered according to their posterior probability for reverse inference, 4 terms were related to pain processing (“painful”, “nociceptive”, “noxious”, and “pain”). In contrast, no term was related to interoception (table 5). The highest posterior probability was reported for the term “painful” (posterior probability=0.77), yet also other pain-related terms (“nociceptive”, “pain”, “noxious”) presented a high posterior probability (p =>0.70). Together, these results indicate that this region presents a high probability to be specifically involved in pain processing.

**Table 5:** Reverse inference metrics [*P*(*term* | *activation*)]: p-value, z-score and posterior probability of the first 15 terms (TERM) and for specific terms (post-hoc TERM) obtained from the decoding of the 6mm spherical region of interest (ROI) centered in the mean of the coordinates forming the cluster obtained with coordinate density analysis (CDA) from Dataset #2 (left anterior insula cortex; mean of the coordinates forming the cluster in MNI space: [−36,17,1]). Abbreviations: post.prob. = posterior probability.

| TERM | Reverse inference metrics |  |  |
| --- | --- | --- | --- |
|  | p-value | z-score | post.prob |
| **painful** | 0.0000 | 7.7138 | 0.7687 |
| choose | 0.0000 | 4.6392 | 0.7592 |
| negative feedback | 0.0003 | 3.6112 | 0.7269 |
| sensation | 0.0001 | 4.0265 | 0.7211 |
| distress | 0.0003 | 3.6329 | 0.7209 |
| **nociceptive** | 0.0009 | 3.3282 | 0.7200 |
| articulatory | 0.0003 | 3.5852 | 0.7186 |
| **noxious** | 0.0005 | 3.4916 | 0.7139 |
| stop signal | 0.0005 | 3.4868 | 0.7074 |
| signal task | 0.0008 | 3.3560 | 0.7070 |
| stop | 0.0000 | 4.1914 | 0.7049 |
| **pain** | 0.0000 | 7.3244 | 0.7037 |
| congruency | 0.0020 | 3.0845 | 0.6994 |
| intense | 0.0033 | 2.9371 | 0.6984 |
| regulatory | 0.0007 | 3.7330 | 0.6960 |
| **post-hoc TERM** |  |  |  |
| *autonomic | 0.1074 | 1.6095 | 0.6152 |
| *interoceptive | 0.2952 | 1.0466 | 0.5945 |

In line with the role of the anterior insula in autonomic and interoceptive domains, the terms “autonomic” and “interoceptive” showed significant results for the forward inference (zscore respectively of 6.75 and 6.93) but not for the reverse inference (table 5). This indicates that the identified region presents consistent associations with interoceptive/autonomic processes, although it is not selectively activated by these processes, as meta-analytically indicated for pain-related processes.

## 4. Discussion

The present study employed two different CBMA approaches (GingerALE and CDA) to determine whether chronic pain disorders are characterized by neurofunctional dysregulations during experimental pain induction. In contrast to two recent neuroimaging meta-analyses, which, surprisingly, did not reveal convergent functional alterations in chronic pain disorders (Tanasescu et al., 2016; Xu et al., 2021), the present study shows robust evidence for altered neurofunctional processing in the left anterior insular cortex of chronic pain patients during experimental pain induction. Notably, functional abnormalities in the left anterior insula cortex were robustly confirmed on Dataset #1 with both ALE and CDA approaches, although with subtle differences, and on Dataset #2 with CDA. Remarkably, the application of CDA on Dataset #2 indicated that the dysfunction of this region in chronic pain patients might be present as either an increased or a decreased activity in comparison to control individuals.

Functional characterization of the identified left insula region employing an unbiased approach (combining a meta-analytic co-activation map and an intrinsic connectivity approach) revealed that this area represents a core node in a functionally integrated network encompassing the bilateral insula cortex, and the anterior cingulate and midcingulate cortex. Moreover, via reverse inference (Poldrack, 2006, 2011), we demonstrated that this anterior insula region has a high probability of being specifically involved in pain processing, with rather unspecific involvement in related processes of interoception.

### 4.1 Anterior insula cortex

The multidimensionality of pain experience (sensory-discriminative, cognitive, and affective-emotional) (Melzack and Casey, 1968) is supported by two main ascending systems: the spino-thalamic cortical pathway and the spino-parabrachial-amygdalar pathway. After the thalamic relay, the spinothalamic cortical route mainly targets the middle cingulate cortex, the posterior insula cortex, and the medial parietal operculum, and, to a less degree, the primary sensory and motor areas (Dum et al., 2009; Lenz et al., 2010). In particular, the posterior insula represents the sensory features of the nociceptive inputs (Frot et al., 2007; Segerdahl et al., 2015; Craig, 2014), while the midcingulate cortex is involved in the processing of negative affects of pain experience (Vogt, 2005) and in the reflexive spatial orientation towards salient sensory stimuli (Vogt, 2009). The spino-parabrachial-amygdalar pathway targets the limbic system through direct projections to the amygdala, playing a major role in the processing of the autonomic and affective responses to pain. In the anterior insula cortex, the sensory information coming from the posterior insula cortex and the affective and autonomic inputs coming from the amygdala are integrated, possibly creating the experience of pain (Bastuji et al., 2018; Frot et al., 2014).

The dysfunction of the anterior insula cortex suggests, therefore, an abnormal and generalized experience of pain processing in chronic pain patients.

In agreement, almost all the studies (7 out of 8) contributing to the convergent cluster of functional abnormality in the left anterior insula cortex compared patients and control participants for the experience (perceived) of pain and not for the intensity of the nociceptive stimulation. Moreover, the studies contributing to the identified cluster induced experimental nociception in as well outside of stimulation site affected by chronic pain, supporting the hypothesis of a generalized dysfunction of acute pain processing in chronic patients, as suggested by previous clinical studies (Staud and Domingo, 2001; Petersel et al., 2011; Chalaye et al., 2012; Goubert et al., 2017).

Notably, the anterior insula cortex plays a complex role in processes directly or indirectly related to the acute and chronic pain experience, including pain empathy (Fallon et al., 2020; Xu et al., 2020b; Zhou et al., 2020), interoception and salience processing (Yao et al., 2018; Li et al., 2018) as well as emotional experience (Gogolla, 2017).

To further disentangle the specific role of the anterior insula region that exhibited neurofunctional dysregulations in chronic pain, a series of independent and meta-analytic functional characterization strategies was utilized.

First, we characterized the corresponding functional network of this region, which consequently may be affected in chronic pain. Determining the co-activation and intrinsic connectivity networks of the identified region in the left anterior insula cortex revealed that it represents a core node of a bilateral network encompassing anterior insular and adjacent ventrolateral prefrontal regions as well as anterior cingulate and midcingulate regions. This network clearly resembles the rs-fMRI “salience” network, anchored in the anterior insula and the midcingulate cortex (Seeley et al., 2007), which plays an important role in the regulation of interoceptive versus external guided attention (Xin et al., 2021; Yao et al., 2018) and the guidance of flexible behavior (Seeley et al., 2007; Menon and Uddin, 2010). Abnormalities in this network have been repeatedly observed in chronic pain (Hemington et al., 2016; Cauda et al., 2009; Wu et al., 2013) and neuropsychiatric conditions, including depression (Manoliu et al., 2014; Shao et al., 2018), anxiety (Geng et al., 2016) or addiction (Klugah-Brown et al., 2021, 2020) which are commonly found to be co-morbid in chronic pain (Bair et al., 2003; Finan and Smith, 2013). These results, therefore, support the hypothesis that chronic pain patients might present an impairment of the salience network and, possibly, of its main functions.

Second, functional decoding via a meta-analytic database (Yarkoni et al., 2011) revealed a high probability for this region to be specifically involved in pain-related processes (terms referred to pain processing: “painful”, “nociceptive”, “noxious”, “pain”), with the term “painful” presenting the highest posterior probability (0.77). Further steps revealed that, although this region is consistently associated with related processes such as interoception and autonomic regulation (as showed by the forward inference metrics), it is not specifically associated with these processes. Together the meta-analytic decoding results suggest that this region, with high probability, is specifically involved in pain processing, yet it plays a broader role in interoceptive/autonomic processes.

### 4.2. Previous meta-analysis

Early neuroimaging meta-analyses performed on chronic pain, although reporting interesting and plausible findings, suffered from a bias towards false-positive results since they employed previous ALE versions, which presented main drawbacks (Eickhoff et al., 2017). However, two robust recent meta-analyses conducted with LocaleALE (Tanasescu et al., 2016) and with the GingerALE updated version (Xu et al., 2021) did not determine robust neurofunctional differences between chronic pain patients and control individuals during experimental pain induction. The null findings in the previous studies (Tanasescu et al., 2016; Xu et al., 2021) might be explained by study selection criteria. In respect to Tanasescu et al. (2016), we enriched the selection of our meta-analysis with other recent investigations and, more importantly, with investigations on irritable bowel syndrome, a visceral chronic pain, not selected in this work. This last observation is critical also in consideration that all the selected studies on irritable bowel syndrome contributed, in our study, to the identified cluster in the left anterior insula cortex. As for the work of Xu et al. (2021), we selected a slightly different dataset of studies. First, we only selected studies investigating chronic pain conditions; second, we excluded chronic pain and pain conditions clearly related to direct central insults (e.g., traumatic brain injury, brain tumors). Moreover, we included in our selection only independent experiments (i.e., we excluded results from similar experiments in the same population) to avoid the problem of the correlation of the reported coordinates (Tanasescu et al., 2016).

### 4.3. Differences in the methods of meta-analysis

Due to the relatively low number of selected papers (14 studies reporting increased activity, 16 studies reporting increased and/or decreased activity), we decided to employ two different CBMA methods: the GingerALE and CDA.

GingerALE is a voxel-wise analysis that requires a stringent threshold (family-wise error) to control type I error. It identifies significant clusters testing the null hypothesis that the peak coordinates are uniformly distributed across the brain. On the other side, CDA estimates the density of reported coordinates from independent studies and uses a model based approach to compute the probability of that, or higher, density under the null hypothesis of uniformly random coordinates. It does this analytically, and with only one parameter (the human grey matter volume).

Being a cluster-wise analysis, less conservative methods can be used, such as the false cluster discovery rate.

## Limitations

This study presents several general and specific limitations that suggest caution in the interpretation of the results. General limitations are related to the common issues of the metaanalysis and of the meta-analysis in neuroimaging. In particular, the well-known problem of publications bias (or file drawer problem), or the tendency to publish studies with significant findings and the less likely probability that studies failing to reject the null hypothesis are published (Ioannidis et al., 2014). This problem is exacerbated in CBMA methods, which are not sensitive to non-significant results also when they are employed in the meta-analysis (Rottschy et al., 2012).

As a specific limitation of this work, we acknowledge the relatively low number of included studies, a drawback of the strict selection criteria (Dataset #1 = 14 studies and Dataset #2 = 16 studies). However, the robustness of our findings are supported by similar results obtained with two different approaches, the GingeALE and the CDA method. Notably, although for GingeALE are recommended at least 17-20 studies (Eickhoff et al., 2016) to obtain robust results, the CDA does not rely on the same recommendations (Tench et al., 2020).

Another important limitation of this work is the massive overrepresentation of women in the selected original studies (83% of chronic pain patients). Therefore, the generalizability of the present findings to men with chronic pain conditions remains to be determined.

It is important to note that in Dataset #2, 2 studies reporting decreased activity in the left anterior insula cortex also contributed, with 6 studies reporting increased activity, to the identified cluster. This might indicate some unknown source of heterogeneity within the selected studies.

## Conclusion

Across two neuroimaging-based meta-analyses, we identified robustly altered neurofunctional processing in patients with chronic pain during nociceptive stimulation. Dysregulations were observed in a cluster located in the left anterior insula, with meta-analytic decoding suggesting that this region is, with high probability, specifically involved in pain processing and not specifically linked to alterations in interoceptive/autonomic processes. On the circuit level, this region represents a core node of a bilateral insular-anterior/mid-cingulate network overlapping with the large-scale salience network.

## Data Availability

Original data will be available upon request.

## References

Albuquerque, R.J., de Leeuw, R., Carlson, C.R., Okeson, J.P., Miller, C.S., Andersen, A.H., 2006. Cerebral activation during thermal stimulation of patients who have burning mouth disorder: an fmri study. Pain 122, 223– 234.

Bair, M.J., Robinson, R.L., Katon, W., Kroenke, K., 2003. Depression and pain comorbidity: a literature review. Archives of internal medicine 163, 2433–2445.

Baliki, M.N., Geha, P.Y., Fields, H.L., Apkarian, A.V., 2010. Predicting value of pain and analgesia: nucleus accumbens response to noxious stimuli changes in the presence of chronic pain. Neuron 66, 149–160.

Bastuji, H., Frot, M., Perchet, C., Hagiwara, K., Garcia-Larrea, L., 2018. Convergence of sensory and limbic noxious input into the anterior insula and the emergence of pain from nociception. Scientific reports 8, 1–9.

Bouhassira, D., Moisset, X., Jouet, P., Duboc, H., Coffin, B., Sabate, J.M., 2013. Changes in the modulation of spinal pain processing are related to severity in irritable bowel syndrome. Neurogastroenterology & Motility 25, 623–e468.

Breivik, H., Collett, B., Ventafridda, V., Cohen, R., Gallacher, D., 2006. Survey of chronic pain in europe: prevalence, impact on daily life, and treatment. European journal of pain 10, 287–333.

Brett, M., Johnsrude, I.S., Owen, A.M., 2002. The problem of functional localization in the human brain. Nature reviews neuroscience 3, 243–249.

Buckner, R.L., Krienen, F.M., Castellanos, A., Diaz, J.C., Yeo, B.T., 2011. The organization of the human cerebellum estimated by intrinsic functional connectivity. Journal of neurophysiology 106, 2322–2345.

Burgmer, M., Pfleiderer, B., Maihöfner, C., Gaubitz, M., Wessolleck, E., Heuft, G., Pogatzki-Zahn, E., 2012. Cerebral mechanisms of experimental hyperalgesia in fibromyalgia. European journal of pain 16, 636–647.

Cauda, F., Sacco, K., Duca, S., Cocito, D., D’Agata, F., Geminiani, G.C., Canavero, S., 2009. Altered resting state in diabetic neuropathic pain. PloS one 4, e4542.

Chalaye, P., Goffaux, P., Bourgault, P., Lafrenaye, S., Devroede, G., Watier, A., Marchand, S., 2012. Comparing pain modulation and autonomic responses in fibromyalgia and irritable bowel syndrome patients. The Clinical journal of pain 28, 519–526.

Collins, D.L., Neelin, P., Peters, T.M., Evans, A.C., 1994. Automatic 3d intersubject registration of mr volumetric data in standardized talairach space. Journal of computer assisted tomography 18, 192–205.

Craig, A., 2014. Topographically organized projection to posterior insular cortex from the posterior portion of the ventral medial nucleus in the long-tailed macaque monkey. Journal of Comparative Neurology 522, 36–63.

Dehghan, M., Schmidt-Wilcke, T., Pfleiderer, B., Eickhoff, S.B., Petzke, F., Harris, R.E., Montoya, P., Burgmer, M., 2016. Coordinate-based (ale) metaanalysis of brain activation in patients with fibromyalgia. Human brain mapping 37, 1749–1758.

Denk, F., McMahon, S.B., Tracey, I., 2014. Pain vulnerability: a neurobiological perspective. Nature neuroscience 17, 192.

Derbyshire, S., Jones, A., Creed, F., Starz, T., Meltzer, C., Townsend, D., Peterson, A., Firestone, L., 2002. Cerebral responses to noxious thermal stimulation in chronic low back pain patients and normal controls. Neuroimage 16, 158–168.

Dueñas, M., Ojeda, B., Salazar, A., Mico, J.A., Failde, I., 2016. A review of chronic pain impact on patients, their social environment and the health care system. Journal of pain research 9, 457.

Dum, R.P., Levinthal, D.J., Strick, P.L., 2009. The spinothalamic system targets motor and sensory areas in the cerebral cortex of monkeys. Journal of Neuroscience 29, 14223–14235.

Eickhoff, S.B., Bzdok, D., Laird, A.R., Kurth, F., Fox, P.T., 2012. Activation likelihood estimation meta-analysis revisited. Neuroimage 59, 2349–2361.

Eickhoff, S.B., Laird, A.R., Fox, P.M., Lancaster, J.L., Fox, P.T., 2017. Implementation errors in the GingerALE Software: description and recommendations. Technical Report 38. Wiley Online Library.

Eickhoff, S.B., Laird, A.R., Grefkes, C., Wang, L.E., Zilles, K., Fox, P.T., 2009. Coordinate-based activation likelihood estimation meta-analysis of neuroimaging data: A random-effects approach based on empirical estimates of spatial uncertainty. Human brain mapping 30, 2907–2926.

Eickhoff, S.B., Nichols, T.E., Laird, A.R., Hoffstaedter, F., Amunts, K., Fox, P.T., Bzdok, D., Eickhoff, C.R., 2016. Behavior, sensitivity, and power of activation likelihood estimation characterized by massive empirical simulation. Neuroimage 137, 70–85.

Eickhoff, S.B., Stephan, K.E., Mohlberg, H., Grefkes, C., Fink, G.R., Amunts, K., Zilles, K., 2005. A new spm toolbox for combining probabilistic cytoarchitectonic maps and functional imaging data. Neuroimage 25, 1325–1335.

Eklund, A., Nichols, T.E., Knutsson, H., 2016. Cluster failure: Why fmri inferences for spatial extent have inflated false-positive rates. Proceedings of the national academy of sciences 113, 7900–7905.

Elsenbruch, S., Rosenberger, C., Enck, P., Forsting, M., Schedlowski, M., Gizewski, E.R., 2010. Affective disturbances modulate the neural processing of visceral pain stimuli in irritable bowel syndrome: an fmri study. Gut 59, 489–495.

Evans, A.C., Collins, D.L., Mills, S., Brown, E.D., Kelly, R.L., Peters, T.M., 1993. 3d statistical neuroanatomical models from 305 mri volumes, in: 1993 IEEE conference record nuclear science symposium and medical imaging conference, IEEE. pp. 1813–1817.

Fallon, N., Roberts, C., Stancak, A., 2020. Shared and distinct functional networks for empathy and pain processing: a systematic review and metaanalysis of fmri studies. Social cognitive and affective neuroscience 15, 709–723.

Finan, P.H., Smith, M.T., 2013. The comorbidity of insomnia, chronic pain, and depression: dopamine as a putative mechanism. Sleep medicine reviews 17, 173–183.

Fox, P.T., Laird, A.R., Lancaster, J.L., 2005. Coordinate-based voxel-wise meta-analysis: Dividends of spatial normalization. report of a virtual workshop. Human brain mapping 25, 1.

Friebel, U., Eickhoff, S.B., Lotze, M., 2011. Coordinate-based meta-analysis of experimentally induced and chronic persistent neuropathic pain. Neuroimage 58, 1070–1080.

Friston, K.J., Holmes, A.P., Price, C., Büchel, C., Worsley, K., 1999. Multisubject fmri studies and conjunction analyses. Neuroimage 10, 385–396.

Frot, M., Faillenot, I., Mauguière, F., 2014. Processing of nociceptive input from posterior to anterior insula in humans. Human brain mapping 35, 5486–5499.

Frot, M., Magnin, M., Mauguière, F., Garcia-Larrea, L., 2007. Human sii and posterior insula differently encode thermal laser stimuli. Cerebral cortex 17, 610–620.

Garcia-Larrea, L., Bastuji, H., 2018. Pain and consciousness. Progress in Neuro-Psychopharmacology and Biological Psychiatry 87, 193–199.

Gaskin, D.J., Richard, P., 2012. The economic costs of pain in the united states. The Journal of Pain 13, 715–724.

Geng, H., Li, X., Chen, J., Li, X., Gu, R., 2016. Decreased intra-and intersalience network functional connectivity is related to trait anxiety in adolescents. Frontiers in Behavioral Neuroscience 9, 350.

Gogolla, N., 2017. The insular cortex. Current Biology 27, R580–R586.

Goldberg, D.S., McGee, S.J., 2011. Pain as a global public health priority. BMC public health 11, 1–5.

Goubert, D., Danneels, L., Graven-Nielsen, T., Descheemaeker, F., Coppieters, I., Meeus, M., 2017. Differences in pain processing between patients with chronic low back pain, recurrent low back pain and fibromyalgia. Pain Physician 20, 307–318.

Gracely, R.H., Petzke, F., Wolf, J.M., Clauw, D.J., 2002. Functional magnetic resonance imaging evidence of augmented pain processing in fibromyalgia. Arthritis & Rheumatism 46, 1333–1343.

Guleria, A., Karyampudi, A., Singh, R., Khetrapal, C.L., Verma, A., Ghoshal, U.C., Kumar, D., 2017. Mapping of brain activations to rectal balloon distension stimuli in male patients with irritable bowel syndrome using functional magnetic resonance imaging. Journal of neurogastroenterology and motility 23, 415.

Hampson, J.P., Reed, B.D., Clauw, D.J., Bhavsar, R., Gracely, R.H., Haefner, H.K., Harris, R.E., 2013. Augmented central pain processing in vulvodynia. The Journal of Pain 14, 579–589.

Hemington, K.S., Wu, Q., Kucyi, A., Inman, R.D., Davis, K.D., 2016. Abnormal cross-network functional connectivity in chronic pain and its association with clinical symptoms. Brain Structure and Function 221, 4203–4219.

Ioannidis, J.P., 2005. Why most published research findings are false. PLoS medicine 2, e124.

Ioannidis, J.P., Munafo, M.R., Fusar-Poli, P., Nosek, B.A., David, S.P., 2014. Publication and other reporting biases in cognitive sciences: detection, prevalence, and prevention. Trends in cognitive sciences 18, 235–241.

Jackson, T., Thomas, S., Stabile, V., Shotwell, M., Han, X., McQueen, K., 2016. A systematic review and meta-analysis of the global burden of chronic pain without clear etiology in low-and middle-income countries: trends in heterogeneous data and a proposal for new assessment methods. Anesthesia & Analgesia 123, 739–748.

Jakobs, O., Langner, R., Caspers, S., Roski, C., Cieslik, E.C., Zilles, K., Laird, A.R., Fox, P.T., Eickhoff, S.B., 2012. Across-study and within-subject functional connectivity of a right temporo-parietal junction subregion involved in stimulus–context integration. Neuroimage 60, 2389–2398.

Jensen, K.B., Kosek, E., Petzke, F., Carville, S., Fransson, P., Marcus, H., Williams, S.C., Choy, E., Giesecke, T., Mainguy, Y., et al., 2009. Evidence of dysfunctional pain inhibition in fibromyalgia reflected in racc during provoked pain. PAIN® 144, 95–100.

Jensen, K.B., Regenbogen, C., Ohse, M.C., Frasnelli, J., Freiherr, J., Lundström, J.N., 2016. Brain activations during pain: a neuroimaging metaanalysis of patients with pain and healthy controls. Pain 157, 1279–1286.

Kim, S.H., Lee, Y., Lee, S., Mun, C.W., 2013. Evaluation of the effectiveness of pregabalin in alleviating pain associated with fibromyalgia: using functional magnetic resonance imaging study. PloS one 8, e74099.

Klugah-Brown, B., Di, X., Zweerings, J., Mathiak, K., Becker, B., Biswal, B., 2020. Common and separable neural alterations in substance use disorders: A coordinate-based meta-analyses of functional neuroimaging studies in humans. Human brain mapping 41, 4459–4477.

Klugah-Brown, B., Zhou, X., Pradhan, B.K., Zweerings, J., Mathiak, K., Biswal, B., Becker, B., 2021. Common neurofunctional dysregulations characterize obsessive–compulsive, substance use, and gaming disorders—an activation likelihood meta-analysis of functional imaging studies. Addiction Biology, e12997.

Kuner, R., Flor, H., 2017. Structural plasticity and reorganisation in chronic pain. Nature Reviews Neuroscience 18, 20.

Laird, A.R., Robinson, J.L., McMillan, K.M., Tordesillas-Gutiérrez, D., Moran, S.T., Gonzales, S.M., Ray, K.L., Franklin, C., Glahn, D.C., Fox, P.T., et al., 2010. Comparison of the disparity between talairach and mni coordinates in functional neuroimaging data: validation of the lancaster transform. Neuroimage 51, 677–683.

Lancaster, J.L., Tordesillas-Gutiérrez, D., Martinez, M., Salinas, F., Evans, A., Zilles, K., Mazziotta, J.C., Fox, P.T., 2007. Bias between mni and talairach coordinates analyzed using the icbm-152 brain template. Human brain mapping 28, 1194–1205.

Lee, J.J., Kim, H.J., Čeko, M., Park, B.y., Lee, S.A., Park, H., Roy, M., Kim, S.G., Wager, T.D., Woo, C.W., 2021. A neuroimaging biomarker for sustained experimental and clinical pain. Nature Medicine 27, 174–182.

Lenz, F.A., Casey, K.L., Jones, E.G., Willis, W.D., 2010. The human pain system: experimental and clinical perspectives. Cambridge University Press.

Li, J., Xu, L., Zheng, X., Fu, M., Zhou, F., Xu, X., Ma, X., Li, K., Kendrick, K.M., Becker, B., 2018. Common and dissociable contributions of alexithymia and autism to domain-specific interoceptive dysregulations–a dimensional neuroimaging approach. bioRxiv, 432971.

Manoliu, A., Meng, C., Brandl, F., Doll, A., Tahmasian, M., Scherr, M., Schwerthöffer, D., Zimmer, C., Förstl, H., Bäuml, J., et al., 2014. Insular dysfunction within the salience network is associated with severity of symptoms and aberrant inter-network connectivity in major depressive disorder. Frontiers in human neuroscience 7, 930.

Melzack, R., Casey, K.L., 1968. Sensory, motivational, and central control determinants of pain: a new conceptual model. The skin senses 1, 423–43.

Menon, V., Uddin, L.Q., 2010. Saliency, switching, attention and control: a network model of insula function. Brain structure and function 214, 655– 667.

Moher, D., Liberati, A., Tetzlaff, J., Altman, D.G., Group, P., et al., 2009. Preferred reporting items for systematic reviews and meta-analyses: the prisma statement. PLoS med 6, e1000097.

Nichols, T., Brett, M., Andersson, J., Wager, T., Poline, J.B., 2005. Valid conjunction inference with the minimum statistic. Neuroimage 25, 653–660.

Petersel, D.L., Dror, V., Cheung, R., 2011. Central amplification and fibromyalgia: disorder of pain processing. Journal of neuroscience research 89, 29–34.

Poldrack, R.A., 2006. Can cognitive processes be inferred from neuroimaging data? sTrends in cognitive sciences 10, 59–63.

Poldrack, R.A., 2011. Inferring mental states from neuroimaging data: from reverse inference to large-scale decoding. Neuron 72, 692–697.

Poldrack, R.A., Baker, C.I., Durnez, J., Gorgolewski, K.J., Matthews, P.M., Munafó, M.R., Nichols, T.E., Poline, J.B., Vul, E., Yarkoni, T., 2017. Scanning the horizon: towards transparent and reproducible neuroimaging research. Nature reviews neuroscience 18, 115.

Pujol, J., López-Solà, M., Ortiz, H., Vilanova, J.C., Harrison, B.J., Yücel, M., Soriano-Mas, C., Cardoner, N., Deus, J., 2009. Mapping brain response to pain in fibromyalgia patients using temporal analysis of fmri. PloS one 4, e5224.

Reid, K.J., Harker, J., Bala, M.M., Truyers, C., Kellen, E., Bekkering, G.E., Kleijnen, J., 2011. Epidemiology of chronic non-cancer pain in europe: narrative review of prevalence, pain treatments and pain impact. Current medical research and opinion 27, 449–462.

Rice, A.S., Smith, B.H., Blyth, F.M., 2016. Pain and the global burden of disease. Pain 157, 791–796.

Rodriguez-Raecke, R., Ihle, K., Ritter, C., Muhtz, C., Otte, C., May, A., 2014. Neuronal differences between chronic low back pain and depression regarding long-term habituation to pain. European Journal of Pain 18, 701–711.

Rottschy, C., Caspers, S., Roski, C., Reetz, K., Dogan, I., Schulz, J., Zilles, K., Laird, A.R., Fox, P.T., Eickhoff, S.B., 2013. Differentiated parietal connectivity of frontal regions for “ what” and “ where” memory. Brain Structure and function 218, 1551–1567.

Rottschy, C., Langner, R., Dogan, I., Reetz, K., Laird, A.R., Schulz, J.B., Fox, P.T., Eickhoff, S.B., 2012. Modelling neural correlates of working memory: a coordinate-based meta-analysis. Neuroimage 60, 830–846.

Schnellbächer, G.J., Hoffstaedter, F., Eickhoff, S.B., Caspers, S., Nicklockschat, T., Fox, P.T., Laird, A.R., Schulz, J.B., Reetz, K., Dogan, I., 2020. Functional characterization of atrophy patterns related to cognitive impairment. Frontiers in neurology 11, 18.

Schreiber, K.L., Loggia, M.L., Kim, J., Cahalan, C.M., Napadow, V., Edwards, R.R., 2017. Painful after-sensations in fibromyalgia are linked to catastrophizing and differences in brain response in the medial temporal lobe. The Journal of Pain 18, 855–867.

Seeley, W.W., Menon, V., Schatzberg, A.F., Keller, J., Glover, G.H., Kenna, H., Reiss, A.L., Greicius, M.D., 2007. Dissociable intrinsic connectivity networks for salience processing and executive control. Journal of Neuroscience 27, 2349–2356.

Segerdahl, A.R., Mezue, M., Okell, T.W., Farrar, J.T., Tracey, I., 2015. The dorsal posterior insula subserves a fundamental role in human pain. Nature neuroscience 18, 499–500.

Seifert, F., Maihöfner, C., 2011. Brain activity associated with pain, hyperalgesia and allodynia: an ale meta-analysis. Journal of neural transmission 118, 1139–1154.

Shao, J., Meng, C., Tahmasian, M., Brandl, F., Yang, Q., Luo, G., Luo, C., Yao, D., Gao, L., Riedl, V., et al., 2018. Common and distinct changes of default mode and salience network in schizophrenia and major depression. Brain imaging and behavior 12, 1708–1719.

Simons, L.E., Moulton, E.A., Linnman, C., Carpino, E., Becerra, L., Borsook, D., 2014. The human amygdala and pain: evidence from neuroimaging. Human brain mapping 35, 527–538.

Staud, R., Domingo, M., 2001. Evidence for abnormal pain processing in fibromyalgia syndrome. Pain Medicine 2, 208–215.

Talairach, J., 1988. Co-planar stereotaxic atlas of the human brain-3-dimensional proportional system. An approach to cerebral imaging.

Tan, L.L., Pelzer, P., Heinl, C., Tang, W., Gangadharan, V., Flor, H., Sprengel, R., Kuner, T., Kuner, R., 2017. A pathway from midcingulate cortex to posterior insula gates nociceptive hypersensitivity. Nature neuroscience 20, 1591.

Tanasescu, R., Cottam, W.J., Condon, L., Tench, C.R., Auer, D.P., 2016. Functional reorganisation in chronic pain and neural correlates of pain sensitisation: a coordinate based meta-analysis of 266 cutaneous pain fmri studies. Neuroscience & Biobehavioral Reviews 68, 120–133.

Tench, C., Tanasescu, R., Constantinescu, C.S., Auer, D.P., Cottam, W., 2020. Coordinate density analysis of neuroimaging studies. bioRxiv.

Tench, C.R., Tanasescu, R., Auer, D.P., Constantinescu, C.S., 2013. Coordinate based meta-analysis of functional neuroimaging data; false discovery control and diagnostics. PloS one 8, e70143.

Tillisch, K., Mayer, E.A., Labus, J.S., 2011. Quantitative meta-analysis identifies brain regions activated during rectal distension in irritable bowel syndrome. Gastroenterology 140, 91–100.

Treede, R.D., Rief, W., Barke, A., Aziz, Q., Bennett, M.I., Benoliel, R., Cohen, M., Evers, S., Finnerup, N.B., First, M.B., et al., 2019. Chronic pain as a symptom or a disease: the iasp classification of chronic pain for the international classification of diseases (icd-11). Pain 160, 19–27.

Turkeltaub, P.E., Eden, G.F., Jones, K.M., Zeffiro, T.A., 2002. Meta-analysis of the functional neuroanatomy of single-word reading: method and validation. Neuroimage 16, 765–780.

Vachon-Presseau, E., Tétreault, P., Petre, B., Huang, L., Berger, S.E., Torbey, S., Baria, A.T., Mansour, A.R., Hashmi, J.A., Griffith, J.W., et al., 2016. Corticolimbic anatomical characteristics predetermine risk for chronic pain. Brain 139, 1958–1970.

Vartiainen, N., Kallio-Laine, K., Hlushchuk, Y., Kirveskari, E., Seppänen, M., Autti, H., Jousmäki, V., Forss, N., Kalso, E., Hari, R., 2009. Changes in brain function and morphology in patients with recurring herpes simplex virus infections and chronic pain. PAIN® 144, 200–208.

Verne, G.N., Himes, N.C., Robinson, M.E., Gopinath, K.S., Briggs, R.W., Crosson, B., Price, D.D., 2003. Central representation of visceral and cutaneous hypersensitivity in the irritable bowel syndrome. PAIN® 103, 99– 110.

Vogt, B.A., 2005. Pain and emotion interactions in subregions of the cingulate gyrus. Nature Reviews Neuroscience 6, 533–544.

Vogt, B.A., 2009. Regions and subregions of the cingulate cortex. Cingulate neurobiology and disease 1.

Wu, Q., Inman, R.D., Davis, K.D., 2013. Neuropathic pain in ankylosing spondylitis: a psychophysics and brain imaging study. Arthritis & Rheumatism 65, 1494–1503.

Xin, F., Zhou, F., Zhou, X., Ma, X., Geng, Y., Zhao, W., Yao, S., Dong, D., Biswal, B.B., Kendrick, K.M., et al., 2021. Oxytocin modulates the intrinsic dynamics between attention-related large-scale networks. Cerebral Cortex 31, 1848–1860.

Xu, A., Larsen, B., Baller, E.B., Scott, J.C., Sharma, V., Adebimpe, A., Basbaum, A.I., Dworkin, R.H., Edwards, R.R., Woolf, C.J., et al., 2020a. Convergent neural representations of experimentally-induced acute pain in healthy volunteers: A large-scale fmri meta-analysis. Neuroscience & biobehavioral reviews 112, 300–323.

Xu, A., Larsen, B., Henn, A., Baller, E.B., Scott, J.C., Sharma, V., Adebimpe, A., Basbaum, A.I., Corder, G., Dworkin, R.H., et al., 2021. Brain responses to noxious stimuli in patients with chronic pain: A systematic review and meta-analysis. JAMA network open 4, e2032236–e2032236.

Xu, X., Dai, J., Liu, C., Chen, Y., Xin, F., Zhou, F., Zhou, X., Huang, Y., Wang, J., Zou, Z., et al., 2020b. Common and disorder-specific neurofunctional markers of dysregulated empathic reactivity in major depression and generalized anxiety disorder. Psychotherapy and psychosomatics 89, 114–116.

Yao, S., Becker, B., Zhao, W., Zhao, Z., Kou, J., Ma, X., Geng, Y., Ren, P., Kendrick, K.M., 2018. Oxytocin modulates attention switching between interoceptive signals and external social cues. Neuropsychopharmacology 43, 294–301.

Yarkoni, T., Poldrack, R.A., Nichols, T.E., Van Essen, D.C., Wager, T.D., 2011. Large-scale automated synthesis of human functional neuroimaging data. Nature methods 8, 665–670.

Yeo, B.T., Krienen, F.M., Sepulcre, J., Sabuncu, M.R., Lashkari, D., Hollinshead, M., Roffman, J.L., Smoller, J.W., Zöllei, L., Polimeni, J.R., et al., 2011. The organization of the human cerebral cortex estimated by intrinsic functional connectivity. Journal of neurophysiology.

Zhou, F., Li, J., Zhao, W., Xu, L., Zheng, X., Fu, M., Yao, S., Kendrick, K.M., Wager, T.D., Becker, B., 2020. Empathic pain evoked by sensory and emotional-communicative cues share common and process-specific neural representations. Elife 9, e56929.

